# Digital applications to support self-management of multimorbidity: A scoping review

**DOI:** 10.1101/2025.04.14.25325811

**Authors:** Lucy Smith, Glenn Simpson, Sian Holt, Hajira Dambha-Miller

## Abstract

**Introduction:** Multimorbidity, defined as the co-occurrence of two or more long-term conditions, is increasing rapidly and poses challenges for healthcare systems. Advances in digital technologies offer solutions by facilitating personalised, scalable care interventions that empower individuals to manage their conditions more effectively. These applications have potential to improve access to care, enhance patient engagement, and support tailored approaches to self-management.

**Objectives:** This scoping review aims to synthesise current evidence on the use of digital applications for self-management in adults with multimorbidity.

**Methods:** A scoping review was conducted, systematically searching PubMed, Web of Science, OVID, CINAHL, EMBASE, and additional manual sources. Boolean operators and targeted key terms were employed to retrieve relevant studies from database inception to 16th January 2024.

**Results:** The search yielded 1,974 articles, of which 31 met the inclusion criteria. Digital applications for self-management in multimorbidity demonstrated high acceptability and varying efficacy. Key benefits included improved communication, symptom tracking, and autonomy. Barriers included privacy concerns, additional patient burden, and engagement challenges. Socio-demographics, self-efficacy, and digital literacy influenced both barriers and facilitators to tool usage. Theoretical models underpinning apps/conceptual models were limited. Older adults and the working-age population were rarely included.

**Conclusion:** The current evidence base does not fully address the needs of older adults with low digital literacy or working-age populations with multimorbidity. Our model highlights the importance of broader contextual mechanisms in digital tool adoption. Future research should prioritise theory-driven tool development tailored to disease clusters and aligned with sociodemographic profiles, health risks, and social care needs. Addressing these gaps could improve self-management and health outcomes for high-risk populations.

**Highlights:**

- Digital applications show promise for supporting self-management of multimorbidity.
- There is limited consideration of how digital applications operate across different patient populations and combinations of diseases.
- There is a paucity of theoretical modelling behind existing digital applications.

## Introduction

Multimorbidity, the coexistence of two or more chronic conditions in an individual, is a growing challenge that places considerable strain on health and social care systems [1]. In England, over one-third of the population now lives with multimorbidity, driving increased demand for health and social care services, while posing complex care delivery challenges [2]. Multimorbidity is associated with higher mortality rates, more frequent hospitalisations, reduced quality of life, and places substantial physical, emotional, and financial burdens on patients and their caregivers [2,3]. Effective self-management, which involves the patient taking an active role in managing symptoms, medications, and lifestyle adjustments, is often promoted as a solution to addressing the challenges of multimorbidity [4]. However, adherence to self-management remains low, with many individuals requiring additional time and support to manage multiple conditions. A reported 436,000 hospital admissions could be avoided if the most vulnerable populations were supported with their self-management [5].

Digital applications, particularly mobile applications, have significant potential for supporting the self-management of multimorbidity. These applications can facilitate remote monitoring, improve communication with care providers, promote treatment plan adherence, and address health and social care needs, etc [7]. Despite their potential, digital interventions have primarily focused on managing single conditions [8], with few addressing the unique complexities of managing multiple, interacting conditions. Existing reviews have either concentrated on older populations or grouped diverse technologies (e.g., web-applications, wearables, telecare) under a single category, limiting insights into the specific role of digital applications for multimorbidity self-management [9].

There is also a need to consider the heterogeneity of people living with multimorbidity. For example, disease clusters associated with multimorbidity vary considerably across age groups, with younger populations experiencing different combinations of conditions compared to older adults, and there are differences arising between ethnic groups and socio-economic backgrounds [1]. Additionally, digital literacy varies by age and socioeconomic status, potentially affecting the usability and efficacy of digital applications in different demographic groups [10].

Addressing these gaps requires a more nuanced understanding of how digital applications can be designed to support the diverse clinical and non-clinical needs of people with multimorbidity. This scoping review aims to synthesise evidence on the use of digital applications for self-management in adults with multimorbidity, with a focus on identifying gaps in knowledge and informing future research and intervention development.

## Methods

A scoping review methodology was used based on the framework developed by Arksey and O’Malley (2005) [11]. This method was chosen as it allows for rapid collation of key evidence in underexplored areas, enabling the synthesis of emerging evidence and trends to identify research gaps and inform future practice, policy and research [12,13].

The review followed the PRISMA-ScR guidelines (Preferred Reporting Items for Systematic Reviews and Meta-Analyses Extension for Scoping Reviews), [14] with the checklist provided in Supplementary Table 1.

The methodology involved six key stages as follows:

### 1. Defining the research question

Based on our preliminary literature searches and the identification of gaps in this field of inquiry, the following research question was addressed:

> *“What is the current evidence on the use of digital applications for self-management in multimorbidity, and what knowledge gaps remain to guide future research?”*

### 2. Identifying relevant studies

The search strategy was developed collaboratively, with a librarian and reviewing previous studies focused on digital applications, self-management, and multimorbidity to inform the initial search terms [9,15]. A reflexive and iterative approach was employed to refine the search strategy. Preliminary searches were conducted on the OVID platform using title and abstract screening to evaluate the relevance of initial terms, enabling adjustments to optimise search outcomes. Both free-text and Medical Subject Headings (MeSH) terms were utilised, and the search terms were reviewed and refined in collaboration with all co-authors. Examples of terms used include:

*multiple chronic conditions OR multimorbidit* AND self-manage* OR self-care OR social care need* OR social support OR digital health OR digital app* OR mobile app* OR mhealth OR website*. The final search terms are presented in Supplementary Table 2.

Systematic electronic searches covered databases including PubMed, Ovid Medline, CINAHL, and Web of Science for articles published from 1946 to December 12, 2024, up to January 16, 2025, were carried out. Manual searching of bibliographies and grey literature searches using BASE, Google, and Google Scholar were also carried out.

#### Inclusion and Exclusion Criteria

Articles were included if they met the following criteria:

- Published in the English language.
- Focused on adult populations with multimorbidity.
- Examined digital applications designed for the self-management of multimorbidity (i.e., two or more long-term conditions).

In line with the exploratory nature of scoping reviews, quality assessment was not a criterion for exclusion. Studies were excluded if they:

- Focused on digital applications developed for a single chronic condition.
- Did not involve self-management.
- Were study protocols, books, or book chapters.
- Lacked full-text availability.

### 3. Selecting relevant studies

All articles identified through the database and grey literature searches were uploaded to the Rayyan citation manager. Duplicates were removed, and titles and abstracts were screened by LS and GS, both of whom have expertise in multimorbidity research. To ensure accuracy, a subset of articles was also screened by SL and SH. Each article was assessed for relevance based on the inclusion criteria. Full-text articles were retrieved for those meeting the initial criteria. Data extraction was conducted independently by LS, GS and SH, with the final selection carried out by LS and GS. Any disagreements were resolved through discussion. In cases of unresolved discrepancies, a third reviewer (SH) made a final decision.

### 4. Data charting

To address the study’s objectives, three data charting tables were used to organise and identify key characteristics of the included studies. These included the: study design, population characteristics, types of long-term conditions, geographic location, the digital application type, self-management targets, theoretical frameworks, outcome measurements, key findings, limitations, and recommendations for future research. Data charting was distributed across the team (GS, LS, SH). This process was iterative, with ongoing communication regarding the relevance of data charting to our research question. Some studies focused solely on digital applications, allowing the full population of all three tables. Others, particularly qualitative studies, explored user experiences and attitudes. Despite this, we deemed it important to include qualitative studies, as they provided valuable insights that enriched our understanding of digital app usage, thereby enhancing the overall comprehensiveness of the review.

### 5. Methods for collating the data

Initially, data were synthesised descriptively through content analysis and frequency counts to summarise study characteristics across all three tables. Extracted data was descriptively interpreted and was structured and categorised in the results. to interpret the data in relation to the research question, identifying key themes and trends. This analysis was led by LS with discussion and review by the wider team for validation.

### 6. Stakeholder feedback

Stakeholder feedback was obtained from our patient representative FD, a South Asian female living with multimorbidity.

## Results

### Screening, inclusion and exclusion of studies

The search of electronic databases and grey literature identified 1,960 articles. After removing 345 duplicates, 1,615 articles were screened. Fourteen citations were also found by hand, making the total number of papers screened 1,629. Taken together, a total of 1,567 articles were excluded, leaving 62 articles eligible for full-text screening and, of these, a final 31 were included (two hand search citations and 29 studies sourced through databases). The PRISMA flowchart summarising the screening process and reasons for exclusion is presented in Table 1.

**Table 1:**
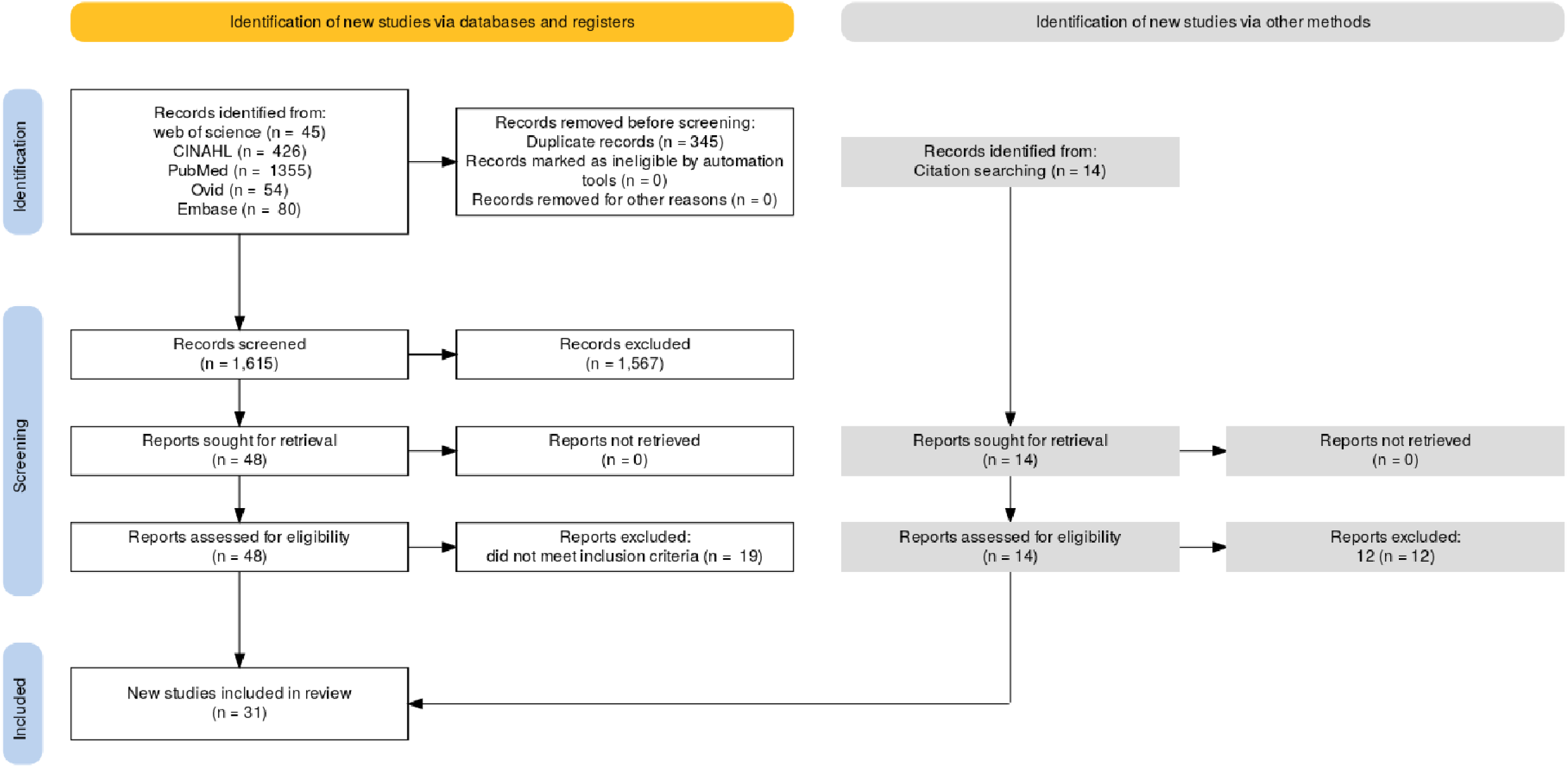
A PRISMA table showing the screening workflow.

### Summary of study characteristics

Data on research methodology are in Supplementary Table 3. All 31 included studies were conducted in high-income countries, with (n = 12) originating from the United States [19,21,23,25,29,31,33,35,41,42,46], (n = 3) from Europe [20,30,36], (n = 4) from Canada [22,28,34,40], (n = 2) from the United Kingdom [17,18], (n = 2) from South Korea [24,45], (n = 2) from China [43,44] (n = 1) from Taiwan [27] and (n = 2) from the Republic of Ireland [32,39].

The timeline of publications (Figure 1) indicates a notable increase in research on multimorbidity and digital applications in recent years.

**Figure 1:**
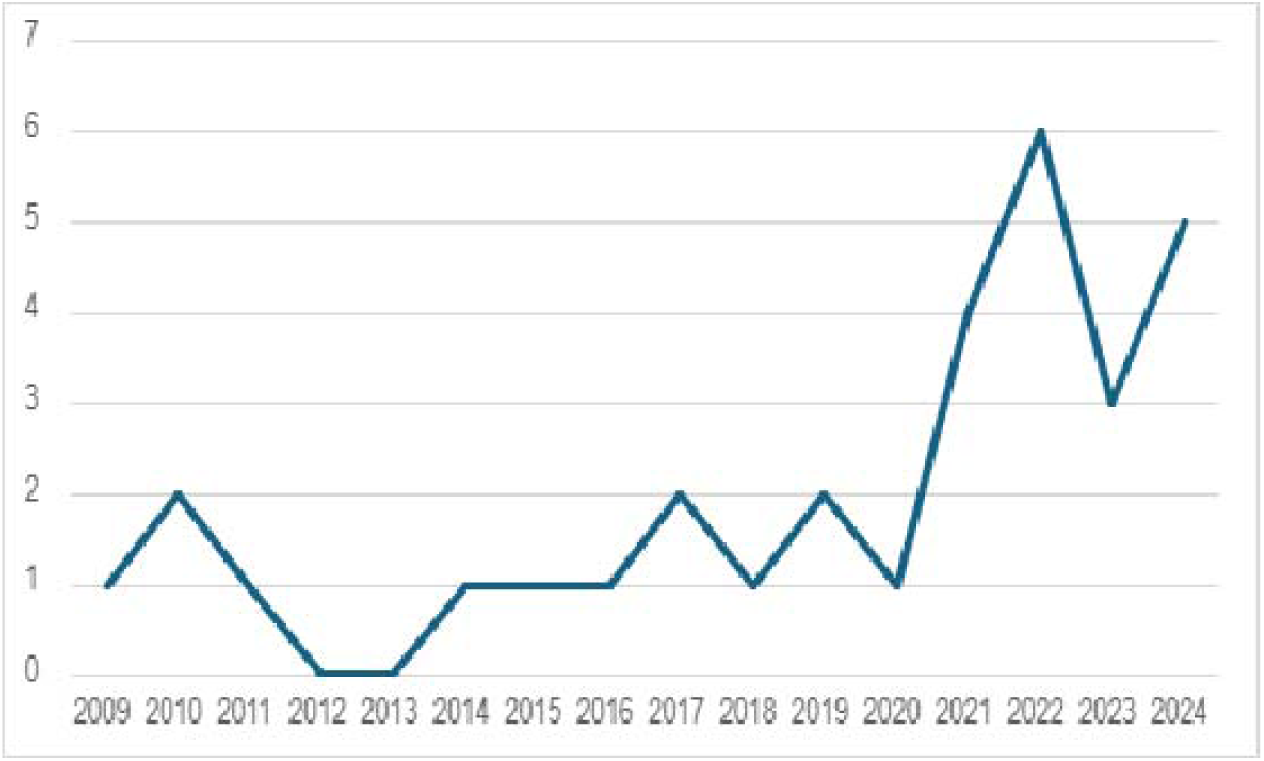
The number of publications of multimorbidity and digital applications by year

Disease representation varied across the studies. Figure 2 illustrates the distribution of six conditions explored in more than four studies. Across all studies, a total of 52 distinct conditions were mentioned (see Supplementary Table 4). Mental health and behavioural conditions were the most frequently referenced conditions (n = 23), followed by cardiovascular disease (n = 11), diabetes (n = 9), hypertension (n = 7), COPD (n = 6) and asthma (n = 4).

**Figure 2:**
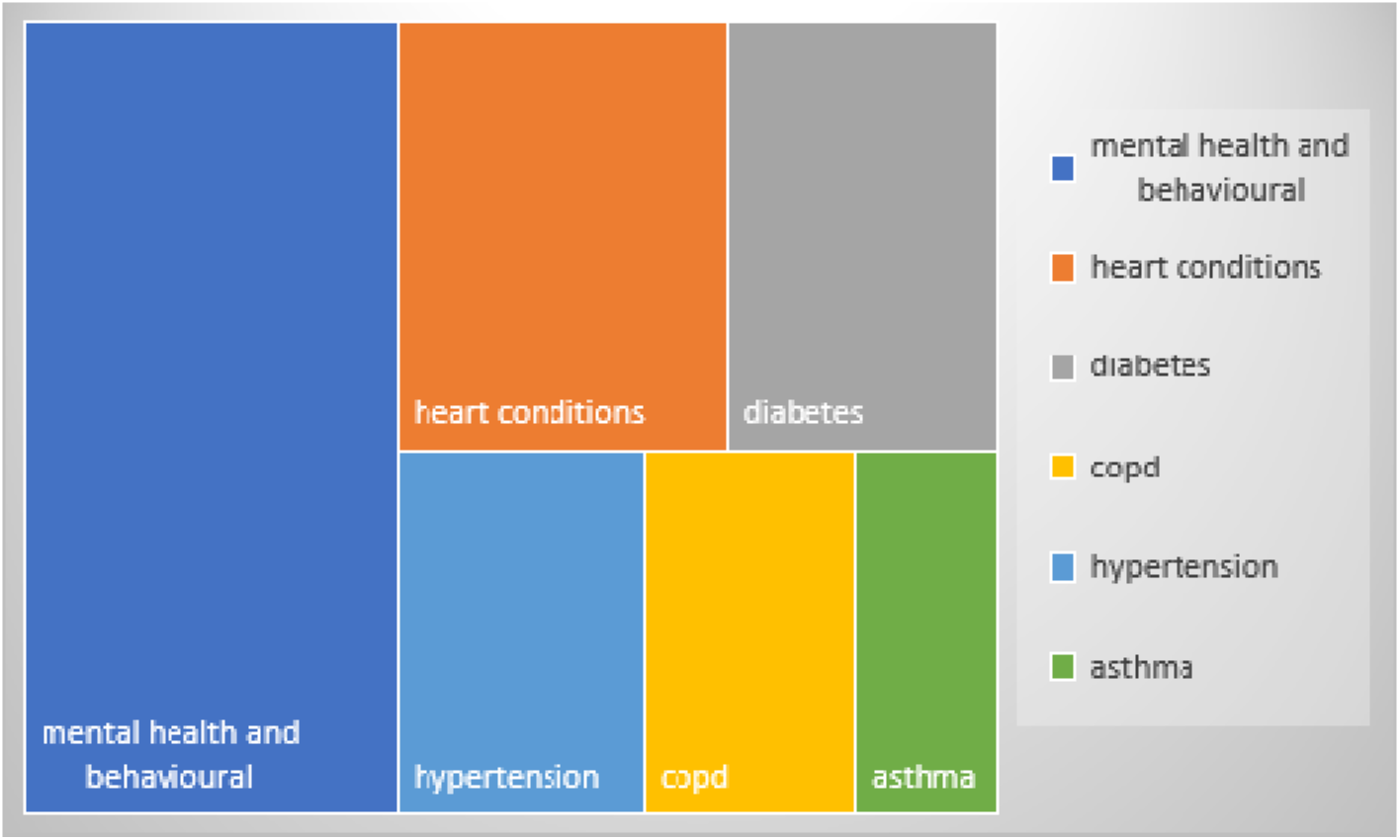
Frequency of conditions within multimorbidity amongst included studies on digital applications

### Objective and categories from included studies

The study objectives were grouped into three main categories.

#### Category 1: App Development and Study methodologies

This category included 17 studies focused on application development or feasibility trials. Methods used included: randomised controlled trials (n = 6) [19,22,23,28,42,45], qualitative studies (n = 2) [26, 34], surveys (n = 1) [17] and combined usability/feasibility trials (n= 8) [7,20,29,30,31,33,38,39].

Qualitative methods were widely employed to gather feedback on issues such as the usability and user experience of applications, including stakeholder workshops, interviews, and think-aloud approaches. Innovative qualitative techniques, such as storyboarding and photo elicitation, were also reported in two studies. All studies recruited participants based on having MLTC, documenting specific diseases in the population retrospectively, and for trial studies, having at least a basic digital proficiency.

#### Category 2: Patient Experiences and Perceptions

Twelve studies explored patient, caregiver, and healthcare professional experiences with digital applications [14,18,24,25,32,35,36,40,41,43,44,46]. Most employed qualitative methods such as interviews and focus groups, while two used surveys. Innovative techniques included storyboarding to depict daily life with MLTC and photo elicitation to document everyday activities.

#### Category 3: Systematic or Scoping Reviews

Two reviews published in 2023 and 2021 explored digital applications for self-management of MLTC. One review [9] included studies from 2009-2019, focusing on older adults (≥60 years) and excluding single-condition interventions, yielding 25 papers. The second review (2010–2020) [37] included 44 studies evaluating self-management support, care coordination, and algorithm-based applications for MLTC management.

### Participant Demographics

Demographic data were available in the 31 included studies. Twenty-five studies (80%) focused on older adults (≥60 years), while six included mixed-age cohorts. None exclusively focused on the working-age population [16–65]. Participants in Category 1 studies were often digitally literate and had higher education levels. Females were overrepresented, with one study exclusively including women [35]. Detailed figures are shown in Supplementary Table 3.

### Type of digital technology used

Table 2 summarises the characteristics of the digital applications and websites in the Theme 1 studies, detailing the type of application, self-management functions, and any relevant theoretical frameworks. Nine studies focused solely on web-based applications, while seven combined web applications with wearable devices to collect biometric, sensory, or clinical data. Ten interventions included a dyadic component, involving healthcare professionals, certified peer mentors, online forums, or caregivers in user interactions or app-generated data.

**Table 2:** The types of applications, content, theoretical underpinnings and self-management strategies for Theme 1 studies.

| Author | Type of app | Self-management functions | Theoretical underpinnings | Self-management strategy |
| --- | --- | --- | --- | --- |
| Balasubramann et al., | Alexa (trialled for at least 2 months). | The tool supported activities of daily living, such as setting alarms to turn off the oven, reminders to do household tasks, suggestions for healthy recipes, info on their illnesses, exercise videos, appointment and medication reminders, social companionship, phone/video call function, helped with sleep. | Not provided in this paper. | Routine management<br>Social support<br>Education. |
| Degroot et al., | Convoy Pal – 12-week intervention using a Routinary Platform, wearable device. | Patients use the app to set health and caregiver needs, and they are then assigned/choose their own goals. They self-monitor symptoms and biometrics from smart watches also feeds in the system. Each week they have a palliative assessment of symptoms, advance care planning, spiritual needs, anticipatory grief, health team needs and social support. The app then responds with messages of support and/or signposts the patient to the correct resource. Patient and carers share the information. Phone calls and Zoom also on the app. | Not provided in this paper. | Goal setting<br>Symptom tracking (self-report and biometrics)<br>Social support<br>Education<br>Dyadic component |
| Dinsmore; Sheng | ProACT: digital app + wearable devices to capture biometrics, accessible to patient, informal carer, formal carer and health professionals. | Patient has wearable device to measure sleep, and biometric applications to measure glucose and blood to monitor vital signs. Alerts are sent if patient has symptoms of clinical concern or has not been on the app. Patient clicks on different 'petals' to set goals, access health tips and entering daily symptoms not trackable by wearable device. Informal carers can access the info and send messages. Formal carers cannot view health data, only wellbeing data, health professionals can view health data. | Not provided in this paper. | Symptom tracking (self-report and biometric)<br>Dyadic component<br>Alert system<br>Education<br>Social support |
| Fortuna | Peer Tech (made using Wellframe)<br>Smartphone app + 1-hour weekly peer support session<br>For patient with a serious mental illness/s + a chronic condition/s. | Certified peer specialist (CPS) allocated, and tablet needed. CPS meets with patients 1 x week for 1 hour to assist with the learning of Emodules targeting psychoeducation and skills training over 3 months. The app supports the learnings from the Emodules, sets personalised self-management goas, allows for medication reminders, with a HIPA A - compliant chat feature. | Based on work from a previous systematic review which advocated use of the integrated illness management and recovery (I-MR). | Education<br>Psychosocial skills<br>Goal setting<br>Health Routine management<br>Social support |
| Gray | ePros portal. | Enables both patients and healthcare providers to identify specific goals and monitor outcomes related to those objectives. Providers use techniques like motivational interviewing, counseling, and health coaching to support patients in managing their health at home between appointments.<br>The Hospital CheckOut feature allows patients to notify their primary care provider when they have been discharged from a hospital visit.<br>The provider portal enables healthcare providers to create care plans in collaboration with patients, outlining the goals they will focus on. Once a goal is included in a patient's Care Plan, it appears in their "My Goal Tracker." This allows patients to track their progress over time either through their mobile device or the patient portal. | The tool is intended to promote a transition to a person-centered care approach by facilitating the entire Goal-Oriented Care process, which includes identifying goals, continuous monitoring, and adjusting goals as needed.<br>The development of the app was guided by the "Fit between Individuals, Task, and Technology" (FITT) framework. | Goal setting<br>Motivational interviewing<br>Routine management<br>Education<br>Social support |
| Gustafon | Eldertree – web-based application. | 12-month trial, web page is a platform which provides information, connection to other elders, motivational services and applications for self-management, to improve quality of life. | Self-determination theory – targets competence, intrinsic motivation and autonomy. | Education<br>Social support<br>Health routine management |
| Kobekyaa | A self and symptom management app – Cancer + other MLTC. | - Make daily health reports<br>- Learn weekly health trends<br>- Schedule reminders through a digital health calendar<br>- Share information<br>- Clear data visualisations<br>- | Grey's self-management framework – needs to be holistic and consider family and patient needs. | Education<br>Routine management<br>Symptom tracker (self-report)<br>Dyadic |
| Lear | Chronic disease management app, manged by Nurse, wearable. | 2-year trial, based on co-ordinated care between patient, nurse, Dr, dietician and exercise specialists.<br>Complete symptom reports – self report and biometric. This could trigger an alert algorithm for the nurse to respond in 1 day. Actions from this – continue with app only, contact DR or hospital. Lifestyle questions completed every 8 weeks, which nurse could use to trigger referral to diet/exercise support team. Public forum, action plan setting, external online resource access and graphical displays of biometric data. | Not referred to in this paper. | Dyadic<br>Symptom tracker (self-report and biometric)<br>Education<br>Social support |
| Massoudi | Personal health record application (PHA) and wearable devices. | Delivers automated, evidence-driven suggestions for physical activity, minimizing the need for continuous oversight and direct engagement from healthcare providers. It monitors users' activity in relation to their set goals and encourages better communication between users and their healthcare providers about physical activity. Users enter data into the app, which also syncs with activity trackers and external services to improve tracking and support. |  | Dyadic<br>Education<br>PA support<br>Goal setting |
| Medinia-Garcia | TenDER technological tool – web-based platform and wearable devices. | The system includes a smartwatch to track physical activity, a sleep monitor to analyze sleep patterns, and a mobile app that displays the collected data. The app also provides self-management tools, such as appointment reminders. | Not referred to in this paper. | Symptom tracking (all from sensory applications)<br>Health Routine management |
| Monahan | Sym Track – tool for monitoring clinically actionable symptoms that are actionable for those with MLTC. | Focus is on 23 non-disease specific clinical symptoms, and separate physical and emotional symptoms for context. Carer report version is also used, for those with informal carers. | Not referred to, but 23 symptoms chosen predict health and economic outcomes independent of disease. | Symptom tracking<br>Dyadic |
| Nambisan | myHESTIA - web based digital app. | The first prototype included 32 trackers, comprising both general trackers (such as for weight and diet, sleep, stress, mood, exercise, and medications) and specific trackers tailored to symptoms or diseases. Each tracker offered different sets of questions or prompts, and patients could choose the ones that were relevant to their conditions. Caregivers were also able to access and add information.<br>The tool featured 9 core elements: (1) trackers, (2) a centralized platform, (3) sense-making applications, (4) gamification, (5) journaling tools, (6) community support, (7) a secure platform, (8) customization options, and (9) communication features, including caregiver involvement. | Not referred to in this paper. | Symptom tracking (self report)<br>Social support<br>Education<br>Dyadic<br>Health routine management |
| Northwood | interRAI Check Up Self Report<br>Digital app to support integrated health and social care for older adults and their care partners. | A tool to help with assessment of need related to cognition, mood, loneliness, pain, instrumental and basic activities of daily living, falls, cardiopulmonary risk, care partner stress, financial trade-offs, health stability, and frailty. Used with care providers to plan health and social care. | Not referred to in this paper. | Symptom tracking<br>Dyadic<br>Social support |
| Shaw | A technological infrastructure to collect and analyse mobile health data from multiple devices available to the public. Platform to combine data from one diet app, and two data transmission applications from Fitbit® and iHealth®. | Track 11 daily health indicators over 4 weeks using devices including steps, physical activity, sleep quantity of sleep, blood pressure, pulse and oxygen saturation, weight and body mass index and fluid intake. | Not referred to in this paper. | Health behaviour tracking (biometric and self-report) |
| Wicks | PatientsLikeMe web-based app online community platform. | Members voluntarily opt to share detailed, measurable data about their symptoms, treatments, and overall health to gain insights from others' experiences and enhance their outcomes. Members can interact through group discussions or private messages. The site's resources are aimed at helping members answer the question, *"Considering my current condition, what is the best possible outcome I can achieve, and what steps do I need to take to reach it?"* | Not referred to in this paper. | Education<br>Social support<br>Dyadic |
| Yoo | Ubiquitous Chronic Disease Care (UCDC) for patients with Type 2 Diabetes, Hypertension and Obesity. | Patients receive phones, glucose blood monitoring equipment, blood pressure equipment and weighing scales. An alarm goes off to take measures, followed by algorithm-defined phrases, determined by clinicians, encouraging words following input related to their health outcomes. Reminders and recommendations also given based on data. Text messages sent to ask about physical activity and weight. Three messages sent a day to give information about diseases and to advise on healthy eating, and exercise methods. Doctors able to access the programme and could send individualised recommendations to their patients. | Not referred to in this paper. | Symptom tracker (biometric and self-report)<br>Education<br>Dyadic<br>Social support |

Social support was the most common feature, present in 10 interventions with human involvement, while six provided digitally generated support via scripted responses, reminders, or reinforcements. Disease education and health improvement strategies were addressed in 11 studies, and symptom tracking was featured in 10, using either self-reported (16 interventions) or biometric (seven interventions) data. Health routine management, including appointment reminders, medication alerts, and sharing information with secondary care professionals, was reported in seven studies.

Stakeholder involvement was consistently integrated throughout the design process, though no study explicitly referenced a theory of change or logic model. Three studies mentioned theoretical frameworks, one was informed by a prior systematic review and another incorporated 23 health outcome variables relevant to various diseases.

### Efficacy of the applications

The findings from the Category 1 studies (app trials) are summarised in Supplementary Table 3 and illustrated in Figure 3. Six studies reported high levels of acceptability and usability. Four studies noted improvements in social well-being and health self-management skills, while two observed enhanced quality of life, better symptom understanding, and reductions in hospitalisations. However, outcome measures varied considerably across studies, with little consistency in the measurement tools used. Negative outcomes were also reported, for example, one study indicated a non-significant decline in quality of life in the intervention group, another found no significant effects between intervention and control groups, one highlighted poor usability satisfaction, and several noted low adherence to device usage.

**Figure 3:**
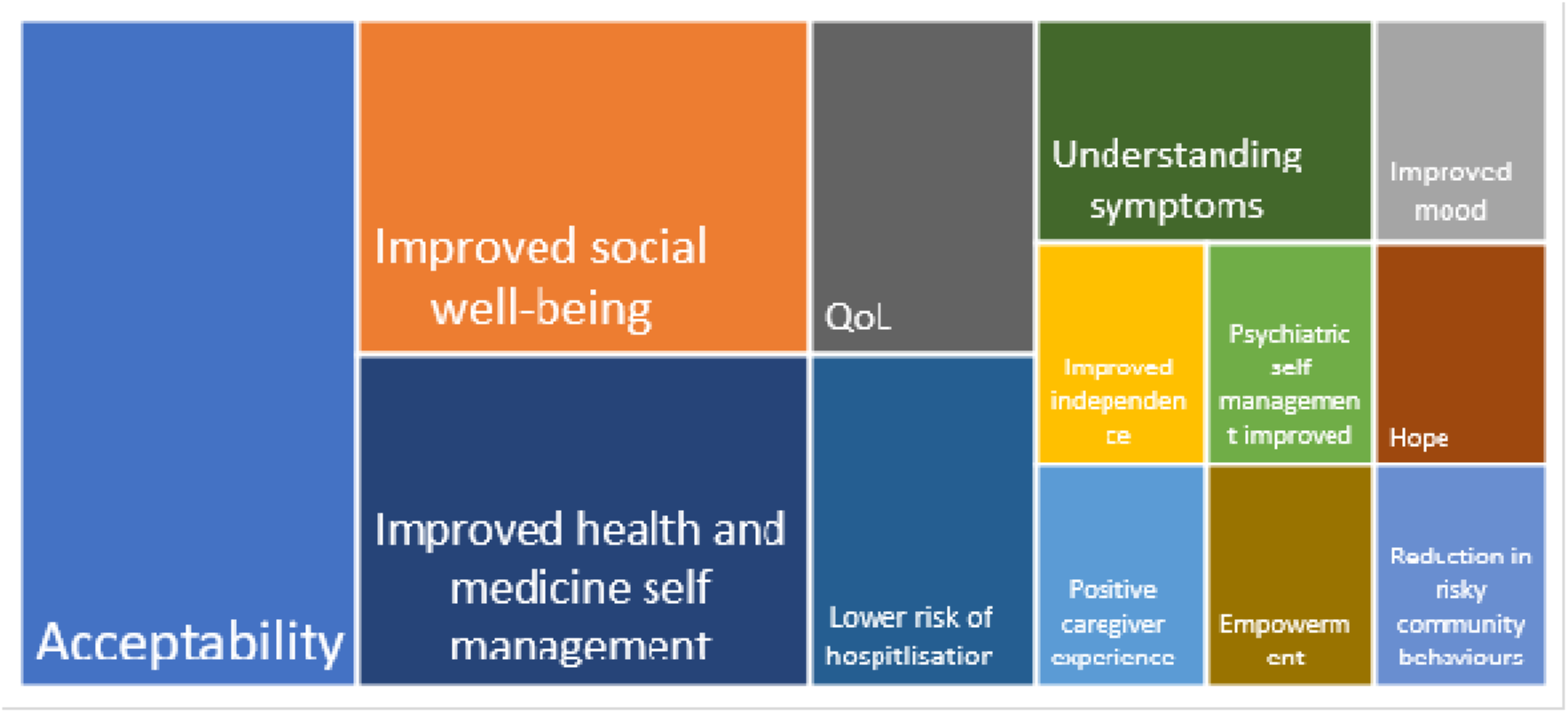
Main effects observed across studies which examined the efficacy of digital applications for the self-management of MLTC

Analysis of findings from Category 2 (qualitative experiences and perceptions) and Category 3 (systematic/scoping reviews), synthesised three overarching themes: barriers, mediators, and facilitators, as shown in Figure 4.

**Figure 4:**
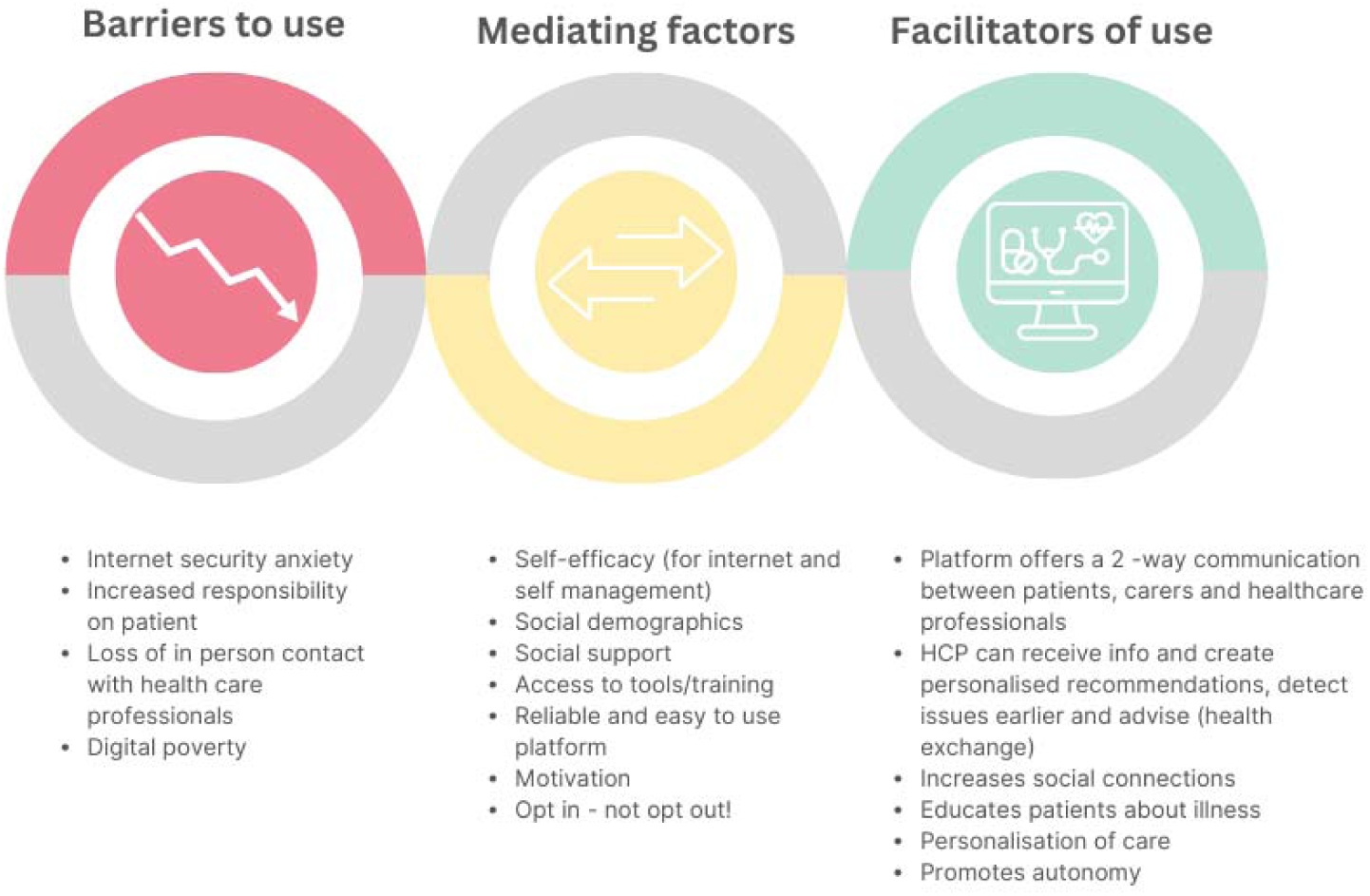
Digital applications and self-management of multiple long-term conditions

### Barriers to Use

The included studies discussed a range of key barriers to digital applications for self-management among people with MLTC. Barriers identified included: concerns related to privacy, security, and data use; the perceived additional burden on patients to manage their health independently; shifting responsibility from traditional healthcare settings; and fears about the potential loss of in-person care.

### Facilitators of Use

A number of facilitators were identified. An important factor discussed in a range of studies were the perceived benefits of digital applications over existing care models and practices. Key benefits identified included improved communication with healthcare professionals, access to personalised recommendations (e.g., early intervention based on symptom feedback), and the ability to tailor applications to individual combinations of conditions and action plans, fostering autonomy and empowerment. Digital applications were also viewed as providing other benefits to patients including facilitating social connectivity with healthcare providers and online patient and advice forums and enabling access to reliable sources of health information and providing skills to support self-management.

### Mediating factors of Use

A number of mediating factors in the use of digital applications were discussed in the studies. An important mediator influencing both barriers and facilitators was socio-demographics. For example, older patients, those with lower education levels or experiencing greater deprivation were more likely to encounter barriers to use, whilst patients with internet access, higher education or digital literacy were more inclined to engage with digital applications.

Self-efficacy also played a key role in accessing and using these applications. Among those with lower self-efficacy digital application use was lower, although there was evidence that training and support from carers or family members could enhance engagement. Further, engagement was reduced if patients felt coerced or pressurised into using digital applications.

### Stakeholder Feedback

Stakeholder input was solicited to assess the research questions and validate the findings of this study. The feedback from our PPIE representative (FD) regarding the research question was:

> *“As a South Asian female living with MLTC, and coming from a low SES background, I appreciate being involved in this important research. I myself live with both mental and physical health conditions, so as a patient - having access to digital apps for my own conditions is very beneficial to me if it will improve my quality of life”*

FD fed back on the conceptual model (Figure 4), emphasizing the need to consider digital poverty as a barrier to use. The conceptual model was amended to incorporate this.

## Discussion

This scoping review aims to synthesise current evidence on the use of digital applications for self-management in adults with multimorbidity. The findings show that within the existing literature, there is limited evidence of how digital applications can be applied to addressing the complexities inherent in managing different combinations of chronic diseases, which is critical to efficacious care in multimorbidity [47,48]. Evidence on this topic focused on high-income states, whilst research on low- and middle-income (LMIC) countries was absent. This may partly reflect the limited rollout of digital applications in LMICs due to a lack of technical systems and delivery infrastructure, patient digital connectivity and implementation costs, etc, although this absence of research is surprising given that there has been significant work in some LMIC countries to leverage digitalise healthcare to increase access to care and improve health outcomes [49–52]. Further, no studies were found that included participants under 18 years old, indicating an important knowledge gap relating to the role of digital applications in supporting self-management of care in multimorbidity across the age spectrum. We found that theoretical frameworks rarely informed application design. While common features such as symptom tracking, social support, and routine management were present, these applications often assumed user motivation based on education about health risks. However, this overlooks key underlying psychosocial factors, such as self-efficacy, which is recognised as playing a crucial role in disease self-management [53].

Our findings are consistent with previous reviews that highlight the lack of attention to the working-age population with multimorbidity and how digital applications can support both the clinical and non-clinical needs of this cohort [1,9]. Although age is an important marker of multimorbidity, over half of the ‘absolute burden of multimorbidity’ is among the working-age population, adding a further layer of complexity for many individuals managing their conditions whilst remaining economically active in the labour market [54]. The absence of this group is concerning given the heterogeneous nature of multimorbidity in this demographic and the need to more effectively support working-age people with chronic conditions to remain economically active and in work for as long as possible [55]. This cohort may benefit from tailored digital interventions, which offer both clinical and non-clinical support (including employment assistance, etc) at an early stage to prevent further deterioration in health or accrual of additional long-term conditions [56, 57]. This will not only have potential to improve care outcomes among multimorbidity patients but also have wider societal benefits in terms of addressing high levels of economic inactivity among working-age populations, which has been a feature of labour markets in many states over the last two decades [58]. The existing evidence base shows that high attrition rates were observed in many app trials, suggesting that results are primarily reflective of digitally engaged participants, who are often white, highly educated, and from more affluent backgrounds [59]. This pattern mirrors the findings of Stone et al. (2020) [60], who noted that the efficacy of digital applications for individuals from lower socioeconomic backgrounds remains under-researched, despite these populations bearing the greatest burden of multimorbidity. The issue of digital inclusion has been described as the “super social determinant of health” [59]. Like previous studies [9, 61], our review found a lack of involvement in research of marginalised populations or those lacking digital literacy. For example, only individuals with internet access and the necessary digital skills were included in trials, which risks exacerbating the digital divide and producing biased and unrepresentative findings. This is particularly concerning as underserved populations, who are more likely to be digitally excluded, face a disproportionate burden of multimorbidity [60,62]. These findings echo the concerns raised by earlier studies on the need for greater efforts to ensure digital health interventions are accessible to all groups, particularly those most affected by chronic conditions [61].

The finding that digital applications were generally well-accepted by participants is also consistent with previous research [63,64]. However, the complexity of self-management across multiple chronic conditions was rarely addressed in the evidence, an issue highlighted in previous work [6]. This contrasts with the call for a more integrated approach to managing multimorbidity. Most digital applications in our review targeted common physical health conditions like diabetes, cardiovascular disease, and hypertension, with only one addressing mental health alongside physical conditions. This finding aligns with recent research by [65,66] a, which highlighted that mental health problems significantly increase the likelihood of physical multimorbidity, underscoring the gap in digital applications for individuals with both chronic physical and mental health conditions. Although psychosocial variables such as quality of life, and social well-being were commonly measured across Category 1 studies through self-reported psychometric questionnaires, only one digital tool directly targeted psychosocial needs [21]. We found that most interventions focused predominantly on biological needs (such as blood pressure and glucose levels, symptom tracking and medication routines) rather than offering practical support for the broader psychosocial needs of individuals with multimorbidity. The biology or clinical dominance in existing applications overlooks the broader challenges faced by patients with multimorbidity, especially when individuals lack the capacity to self-manage [67]. This further highlights the risk of increasing the burden on people with multimorbidity through the use of digital applications without the concomitant provision of appropriate user support. Our review also identified other important potential barriers to the use of digital applications, such as privacy concerns, the burden of managing health digitally, and fears about losing in-person care. These barriers have been identified in earlier research limiting access to digital health applications among particular groups [63,64]. However, few studies have explored these within the context of multimorbidity or outlined specific measures to mitigate potential barriers to digital application take-up among this cohort.

This review has several limitations. While we employed a comprehensive search strategy, the exclusion of telehealth and wearable-only interventions may have restricted the breadth of our findings. Additionally, we limited our inclusion criteria to studies that explicitly referenced multimorbidity and multiple long-term conditions, potentially overlooking studies involving older adults with multimorbidity that did not specifically label their population as such. By focusing on studies involving both digital applications and wearable devices, we may have unintentionally narrowed the scope, potentially omitting insights from research that did not involve these technologies. Furthermore, while stakeholder feedback was integrated into the review, its value would have been enhanced if it had been embedded in the research design from the outset, allowing for a more iterative and informed approach to the review process.

Despite these limitations, the review possesses several notable strengths. We included a range of patient populations, with a particular focus on working-age adults, a group often underrepresented in existing literature on multimorbidity. This focus helps address a significant gap in the current research, as working-age adults with multimorbidity have specific challenges that are less frequently explored. Additionally, we adhered to a rigorous scoping review methodology, ensuring an extensive and systematic search of both grey literature and peer-reviewed publications, which contributed to the comprehensiveness of the findings. This methodical approach strengthens the validity of the review by capturing a broad spectrum of evidence across diverse contexts.

## Conclusion

Digital applications for self-management of multimorbidity continue to be developed, but research remains limited and fragmented, often neglecting the complexities of managing multiple chronic conditions clinically and in relation to non-clinical factors like psychological support and social care. Current evidence does not sufficiently assess the efficacy of these applications for underserved populations, including older adults with low digital literacy and working-age individuals with chronic conditions.

Our conceptual model highlights the importance of considering broader contextual mechanisms influencing digital tool uptake. To move forward, digital self-management interventions must consider psychosocial factors, such as self-efficacy, involve standardised outcome measures and incorporate evidence based on long-term trials. Future research should focus on tailoring interventions to specific multimorbidity clusters and vulnerable populations to ensure greater accessibility and effectiveness, following theory-driven intervention development processes. By addressing these gaps, digital applications can become more accessible, inclusive, and effective in improving the self-management of multimorbidity across diverse groups.

## Acknowledgements

We would like to acknowledge Dr Sridhar Lankey and Dr Neil Singh for their contributions towards the screening process, and Firoza Davies for providing patient feedback on the manuscript.

## Ethical Considerations

Ethical approval was not required.

## Consent to Participate

Not applicable.

## Consent for Publication

Not applicable.

## Data Availability

Data are available from authors at reasonable request.

## Supplementary materials

**Supplementary Table 1:** Preferred Reporting Items for Systematic reviews and Meta-Analyses extension for Scoping Reviews (PRISMA-ScR) Checklist.

| SECTION | ITEM | PRISMA-ScR CHECKLIST ITEM | REPORTED ON PAGE # |
| --- | --- | --- | --- |
| **TITLE** |  |  |  |
| Title | 1 | Identify the report as a scoping review. | Front page of article |
| **ABSTRACT** |  |  |  |
| Structured summary | 2 | Provide a structured summary that includes (as applicable): background, objectives, eligibility criteria, sources of evidence, charting methods, results, and conclusions that relate to the review questions and objectives. | 2 |
| **INTRODUCTION** |  |  |  |
| Rationale | 3 | Describe the rationale for the review in the context of what is already known. Explain why the review questions/objectives lend themselves to a scoping review approach. | 3 |
| Objectives | 4 | Provide an explicit statement of the questions and objectives being addressed with reference to their key elements (e.g., population or participants, concepts, and context) or other relevant key elements used to conceptualize the review questions and/or objectives. | 3-4 |
| **METHODS** |  |  |  |
| Protocol and registration | 5 | Indicate whether a review protocol exists; state if and where it can be accessed (e.g., a Web address); and if available, provide registration information, including the registration number. | No formal review protocol |
| Eligibility criteria | 6 | Specify characteristics of the sources of evidence used as eligibility criteria (e.g., years considered, language, and publication status), and provide a rationale. | 4 |
| Information sources* | 7 | Describe all information sources in the search (e.g., databases with dates of coverage and contact with authors to identify additional sources), as well as the date the most recent search was executed. | 4,26-28 |
| Search | 8 | Present the full electronic search strategy for at least 1 database, including any limits used, such that it could be repeated. | 26-28 |
| Selection of sources of evidence† | 9 | State the process for selecting sources of evidence (i.e., screening and eligibility) included in the scoping review. | 4-5 |
| Data charting process‡ | 10 | Describe the methods of charting data from the included sources of evidence (e.g., calibrated forms or forms that have been tested by the team before their use, and whether data charting was done independently or in duplicate) and any processes for obtaining and confirming data from investigators. | 5 |
| Data items | 11 | List and define all variables for which data were sought and any assumptions and simplifications made. | 4 |
| Critical appraisal of individual sources of evidence§ | 12 | If done, provide a rationale for conducting a critical appraisal of included sources of evidence; describe the methods used and how this information was used in any data synthesis (if appropriate). | Not conducted |
| Synthesis of results | 13 | Describe the methods of handling and summarizing the data that were charted. | 5 |
| **RESULTS** |  |  |  |
| Selection of sources of evidence | 14 | Give numbers of sources of evidence screened, assessed for eligibility, and included in the review, with reasons for exclusions at each stage, ideally using a flow diagram. | 6 |
| Characteristics of sources of evidence | 15 | For each source of evidence, present characteristics for which data were charted and provide the citations. | 29-39 |
| Critical appraisal within sources of evidence | 16 | If done, present data on critical appraisal of included sources of evidence (see item 12). | Not conducted as not part of scoping review methodology |
| Results of individual sources of evidence | 17 | For each included source of evidence, present the relevant data that were charted that relate to the review questions and objectives. | 29-39 |
| Synthesis of results | 18 | Summarize and/or present the charting results as they relate to the review questions and objectives. | 10-12 |
| **DISCUSSION** |  |  |  |
| Summary of evidence | 19 | Summarize the main results (including an overview of concepts, themes, and types of evidence available), link to the review questions and objectives, and consider the relevance to key groups. | 13-17 |
| Limitations | 20 | Discuss the limitations of the scoping review process. | 16-17 |
| Conclusions | 21 | Provide a general interpretation of the results with respect to the review questions and objectives, as well as potential implications and/or next steps. | 17 |
| **FUNDING** |  |  |  |
| Funding | 22 | Describe sources of funding for the included sources of evidence, as well as sources of funding for the scoping review. Describe the role of the funders of the scoping review. | 17 |

JB1 = Joanna Briggs Institute; PRISMA-ScR = Preferred Reporting Items for Systematic reviews and Meta-Analyses extension for Scoping Reviews.
\* Where *sources of evidence* (see second footnote) are compiled from, such as bibliographic databases, social media platforms, and Web sites.
† A more inclusive/heterogeneous term used to account for the different types of evidence or data sources (e.g., quantitative and/or qualitative research, expert opinion, and policy documents) that may be eligible in a scoping review as opposed to only studies. This is not to be confused with *information sources* (see first footnote).
‡ The frameworks by Arksey and O'Malley (6) and Levac and colleagues (7) and the JBI guidance (4, 5) refer to the process of data extraction in a scoping review as data charting.
§ The process of systematically examining research evidence to assess its validity, results, and relevance before using it to inform a decision. This term is used for items 12 and 19 instead of "risk of bias" (which is more applicable to systematic reviews of interventions) to include and acknowledge the various sources of evidence that may be used in a scoping review (e.g., quantitative and/or qualitative research, expert opinion, and policy document).
From: Tricco AC, Lillie E, Zarin W, O'Brien KK, Colquhoun H, Levac D, et al. PRISMA Extension for Scoping Reviews (PRISMA-ScR): Checklist and Explanation. *Ann Intern Med*. 2018;169:467–473. doi: [10.7326/M18-0850](https://doi.org/10.7326/M18-0850).

**Supplementary Table 2:** Summary of search terms.

| MeSH and Free Text Search Terms | Databases | Filters/Refined by | Number of Sources identified |
| --- | --- | --- | --- |
| (multiple chronic conditions or multimorbidit* or multiple long term conditions or multiple long-term conditions or multiple chronic diseases or multiple chronic illnesses or multiple health conditions).mp.<br>(self-manage* or self-manage* or self-care or self care).mp.<br>(digital health or digital app* or mobile app* or mhealth or website or webpage or web-page or smart* or e-health or ehealth or digital technolog* or intervention or tool* or device or platform or remote monitor* or assistive).mp.<br>(social care need* or social support or activities of daily living or ADL or financ* or housing or mobility or bereavement or social net* or residential or community care).mp. | Ovid MEDLINE | Searches from database inception.<br>Language: restricted to the English language.<br>Adults. | **54** |
| (multiple chronic conditions OR multimorbidit* OR multiple long term conditions OR multiple long-term conditions OR multiple chronic diseases OR multiple chronic illnesses OR multiple health conditions ) AND ( self-manage* OR self-manage* OR self-care OR self care ) AND ( digital health OR digital app* OR mobile app* OR mhealth OR website OR webpage OR web-page OR smart* OR e-health OR ehealth OR digital technolog* OR intervention OR tool* OR device OR platform OR remote monitor* OR assistive ) AND ( social care need* OR social support OR activities of daily living OR ADL OR financ* OR housing OR mobility OR bereavement OR social net* OR residential OR community care) | CINAHL (EBSCO) | Searches from database inception.<br>Language: restricted to the English language.<br>Adults. | **426** |
| (multiple chronic conditions or multimorbidit* or multiple long term conditions or multiple long-term conditions or multiple chronic diseases or multiple chronic illnesses or multiple health conditions).mp.<br>(self-manage* or self-manage* or self-care or self care).mp.<br>(digital health or digital app* or mobile app* or mhealth or website or webpage or web-page or smart* or e-health or ehealth or digital technolog* or intervention or tool* or device or platform or remote monitor* or assistive).mp.<br>(social care need* or social support or activities of daily living or ADL or financ* or housing or mobility or bereavement or social net* or residential or community care).mp. | EMBASE | Searches from database inception.<br>Language: restricted to the English language.<br>Adults. | **80** |
| ((multiple chronic conditions OR multimorbidit* OR multiple long term conditions OR multiple long-term conditions OR multiple chronic diseases OR multiple chronic illnesses OR multiple health conditions) AND (self-manage* OR self-manage* OR self-care OR self care)) AND (digital health OR digital app* OR mobile app* OR mhealth OR website OR webpage OR web-page OR smart* OR e-health OR ehealth OR digital technolog* OR intervention OR tool* OR device OR platform OR remote monitor* OR assistive)) AND (social care need* OR social support OR activities of daily living OR ADL OR financ* OR housing OR mobility OR bereavement OR social net* OR residential OR community care) | PubMed | Searches from database inception.<br>Language: restricted to the English language.<br>Adults. | **1,355** |
| ((TS=(multiple chronic conditions OR multimorbidit* OR multiple long term conditions OR multiple long-term conditions OR multiple chronic diseases OR multiple chronic illnesses OR multiple health conditions)) AND TS=(self-manage* OR self-manage* OR self-care OR self care)) AND TS=(digital health OR digital app* OR mobile app* OR mhealth OR website OR webpage OR web-page OR smart* OR e-health OR ehealth OR digital technolog* OR intervention OR tool* OR device OR platform OR remote monitor* OR assistive)) AND TS=(social care need* OR social support OR activities of daily living OR ADL OR financ* OR housing OR mobility OR bereavement OR social net* OR residential OR community care) | Web of Science | Searches from database inception.<br>Language: restricted to the English language.<br>Adults. | **45** |

**Supplementary Table 3:** Study characteristics and findings for all included studies focused on digital applications, MLTC and self-management.

| Author & title | Paper reference number | Study year | Study design | Sample | MLTCs targeted | Aim/s of the study | Country of origin | Findings |
| --- | --- | --- | --- | --- | --- | --- | --- | --- |
| Balasubramanian et al.,<br>Digital personal assistants are smart ways for assistive technology to aid the health and wellbeing of patients and carers. | 17 | 2021 | Service evaluation / market research. | ‘Telephone survey and two focus groups of patients (n = 44) and informal carers (n = 7).<br>The age of the participants ranged from 50 to 90 years.<br>No other demographics reported.’ | ‘Focus groups were for patients with diabetes but most had existing comorbidities.<br>Survey responders had:<br>Diabetes, dementia, Parkinson’s disease, asthma, Behçet’s disease, Cushing’s syndrome, phenylketonuria, liver disorders, low mood, depression, anxiety, dyslexia, cognitive impairment, severe visual impairment with a disability to read and write, chronic knee pain affecting mobility and trauma.’ | ‘To assess the efficacy of the digital tool.’ | UK | ‘Improved health and social wellbeing, reminders about meds helped improve adherence and independence increased, low mood and feeling lonely decreased.’ |
| Cajamarca et al.,<br>Technologies for Managing the Health of Older Adults with Multiple Chronic Conditions: A Systematic Literature Review. | 9 | 2023 | Systematic review. | ‘681 papers extracted from last 10 years, final n – 2.5.’ | Any included. | ‘To describe the technologies that are used to support the management of multimorbidity for older adults.’ | International | ‘Applications and websites focused on communication and connectivity mostly used. Raised challenges of targeting multi-illnesses in one app, as they are still being treated as one illness.’ |
| Cummings et al.,<br>Patient co-design of digital health storytelling applications for multimorbidity: A phenomenological study. | 18 |  | ‘Narrative within a phenomenological approach to inform a process of co-design.’ | ‘One man and four women between the ages of 31 and 53.’ | ‘Borderline personality disorder, chronic migraines, depression and anxiety, early menopause, fibromyalgia, irritable bowel syndrome, myalgic encephalomyelitis (ME), polycystic ovary syndrome, posttraumatic stress disorder, postural tachycardia syndrome, psoriatic arthritis, Raynaud’s disease and relapsing-remitting multiple sclerosis.’ | ‘Using story telling applications to find out what is important in the lives of those with MLTC.’ | Scotland, UK | ‘Participants need to communicate the big picture of their illness, to be able to explain how they experience their illness and how to challenge scepticism from others. Health stories can be used to foster better communication with others about their illness.’ |
| DeGroot et al.,<br>Feasibility of a digital palliative care intervention (Convoy-Pal) | 19 | 2024 | RCT. | ‘31 patients - mean age: 76.3. 15 Caregivers - mean | ‘Two or more self-reported multiple chronic conditions (MCC) including Heart Failure (HF) and score higher | ‘To test the feasibility of delivering the | USA | ‘Good acceptability for the tool, patients reported better social functioning, |
| for older adults with heart failure and multiple chronic conditions and their caregivers: a waitlist randomized control trial. |  |  |  | age: 71.6.’ | than 2 on the Disease Burden/ Morbidity Assessment indicating the presence of MCC that limit activities of daily of living.’ | Convoy-Pal intervention.’ |  | and caregivers improved their positive perceptions of caregiving.’ |
| Dinsmore et al.,<br>ProACT: Person-centred digital integrated care for adults aged 65 years and over, living with multimorbidity. | 20 | 2019 | Mixed methods study. | ‘Mixed methods study consisting of: a qualitative user requirements study.(n=124 stakeholders); co-design workshops and usability testing (n=60 stakeholders); a 12-month longitudinal, action research proof of concept trials (involving n=120 patients and their care networks).’ | None specified. | ‘To assess the efficacy of the digital tool for over 65s with MLTC.’ | Europe – Ireland, Italy and Belgium. | ‘Older patients may be well supported to manage their care needs at home using this app.’ |
| Fortuna et al.,<br>Feasibility, Acceptability, and Preliminary Effectiveness of a Peer-Delivered and Technology Supported.<br>Self-Management Intervention for Older Adults with Serious Mental Illness. | 21 | 2018 | Feasibility study. | ‘Ten adults, age (range 62 to 77, mean 68.8).’ | ‘Diagnosis of schizophrenia, schizoaffective disorder, bipolar disorder, or major depressive disorder, and (4) at least one medical condition defined as cardiovascular disease, obesity, diabetes, chronic obstructive pulmonary disease, chronic pain, hypertension, or high cholesterol.’ | ‘To assess the efficacy of a digital tool for older adults with serious mental illness.’ | USA | ‘PeerTECH showed significant improvements in psychiatric self-management, self-efficacy for managing chronic health conditions, hope, quality of life, medical self-management skills, and empowerment.’ |
| Gray et al.,<br>Assessing the Implementation and Effectiveness of the Electronic Patient-Reported Outcome Tool for Older Adults With Complex Care Needs: Mixed Methods Study. | 22 | 2021 | Randomized controlled trial using a stepped-wedge design and ethnographic case studies. | Six primary care practices, 2 rural, 2 urban, 2 mixed rural. 42 patients. | Arthritis, Asthma, Atrial fibrillation, Cancer Chronic, obstructive pulmonary disease, Congestive heart failure. Diabetes, Enlarged prostate, Epilepsy, Gastroparesis, Hypercholesterolemia. Hypertension, Hypothyroidism, Ischemic heart disease Kidney failure, Macular degeneration, Mental health conditions, Pain, Sleep apnea, Stroke, Urinary retention, Other. | ‘To assess the efficacy of ePRO.’ | Canada | ‘No significant effects observed.’ |
| Gustafson et al.,<br>Effect of an eHealth intervention on older adults' quality of life and health-related outcomes: a randomized clinical trial. | 23 | 2022 | Non blinded randomized controlled trial, 2 arm. | '390 adults age $\geq 65$ with health challenges. Exclusions: long-term care, inability to get out of bed/chair unassisted.' | Not listed. | 'To assess the effects of an eHealth intervention for older adults.' | USA | 'There were no main effects of ElderTree over time. Moderation analyses indicated that among participants with high primary care use, ElderTree (vs. control) led to better trajectories for mental quality of life social support received, social support provided, and depression.' |
| Ha, et al.,<br>Factors Affecting the Acceptability of Technology in Health Care Among Older Korean Adults with Multiple Chronic Conditions: A Cross-Sectional Study Adopting the Senior Technology Acceptance Model. | 24 | 2020 | Survey. | '226 community-dwelling older adults with more than two chronic conditions participated in this study, ages ranging from 66 to 96 years. Mean age 79.' | Hypertension, diabetes, heart disease, thyroid disease, and cancer. | 'This study aimed to investigate the acceptance of technology among older Korean adults with multiple chronic health conditions.' | South Korea | 'Older Korean adults with multiple chronic health conditions scored moderately high for technology acceptance. There were significant differences in technology acceptance according to age, cognitive ability, gerontechnology self-efficacy and facilitating conditions Age and education were significant factors predicting technology acceptance.' |
| Haverhals. et al.,<br>Older adults with multi-morbidity: medication management processes and design implications for personal health applications. | 25 | 2011 | 'A qualitative analysis of a series of individual and group semi-structured interviews with participants who were identified through purposive sampling.' | 32 adult patients and 2 adult family caregivers. | No specific conditions –chronic medical conditions. | 'To understand the medication self-management issues faced by older adults and caregivers and views on digital applications.' | USA | 'The results suggest that PHAs should have the following features to accommodate the management strategies and information preferences of this population: (1) provide links to authoritative and reliable information on side effects, drug interactions, and other medication-related concerns in a way that is clear, concise, and easy to navigate, (2) facilitate communication between patients and doctors and pharmacists through electronic messaging and health information exchange, and (3) provide patients the ability to selectively disclose medication information to different clinicians.' |
| Kobekyaa et al.,<br>A Tailored Self-Management App to Support Older Adults with Cancer and Multi-Morbidities: Development and Usability Testing. | 26 | 2024 | Design thinking model – qualitative. | '18 – 15 older people over 65 who had cancer treated in the last year + 1 other chronic illness which required medication, 3 caregivers.' | Cancer, Asthma, emphysema, chronic bronchitis, or COPD, Arthritis or Rheumatism, Diabetes, Digestive problems, heart problems, depression and anxiety, other. | 'To report on user evaluations of the design of a self-management app to support older adults living with cancer and multimorbidity.' | Canada | 'Participants highlighted the importance of tracking functions to make sense of the information about their symptoms, clear displays, and reminders to mitigate concerns related to polypharmacy.' |
| Kuo et al.,<br>Use of photo diary and focus group to explore needs for digital disease management program among community older adults with chronic disease. | 27 | 2022 | 'Photo diary and focus groups. Photos taken for 3 months of daily activities and aspects of daily living that ppps felt were important to them in relation to managing their illness.' | 'Eleven older adults with multiple metabolic diseases, mean age 72.5, living in a care home.' | CVD, Type 2 diabetes, hypertension. | 'To explore the needs for disease management in activity tracking using photo diary through older adults' subjective perspective.' | Taiwan | 'Five subthemes (regular physical activity, smart management of healthy behaviors, healthy diet, regular daily routine and social connection) were identified.' |
| Lear et al.,<br>Assessment of an Interactive Digital Health-Based Self-management Program to Reduce Hospitalizations Among Patients With Multiple Chronic Diseases: A Randomized Clinical Trial. | 28 | 2021 | 'Single blind, randomized, clinical trial, randomized to receive digital chronic management intervention or normal hospital care.' | '229 patients, mean age = 70.5, 61.6% males.' | Heart diseases, COPD, diabetes, chronic kidney disease. | 'To compare the effect of an internet-based self-management and symptom monitoring program targeted to patients with MLTC with usual care on hospitalizations over a 2-year period.' | Canada | 'The internet CDM group had 25 fewer hospitalizations compared with the usual care group. The intervention group also had 229 fewer in-hospital days compared with the usual care group and social support improved in the intervention group. Fewer participants in the internet CDM vs usual care group had at least 1 hospitalization individuals or experienced the composite outcome of all-cause hospitalization or participants in the internet CDM group had a lower risk of time to first hospitalization.' |
| Massoudi, et al.,<br>A web-based intervention to support increased physical activity among at-risk adults. | 29 | 2010 | Developing a new app. | 'Phase1: 28 consumers (sedentary adults with and without chronic health conditions), 8 health care providers (physicians, nurses, and physical therapists), and 6 personal trainers.<br>Phase 2: 8 participants (5 consumers and 3 health care providers).' | Diabetes, high blood pressure, coronary heart disease, and obesity. | 'To prototype a personal health record application (PHA) that delivers and supports a highly individualized, behaviorally based lifestyle physical activity intervention for sedentary adults.' | USA | 'The PHA was developed to include elements of cognitive and behavioral skills such as goal-setting, self-monitoring, accepting social support, cognitive restructuring, contingency management, decisional balance, and relapse prevention.' |
| Medina-Garcia, et al.,<br>A Technological Tool Aimed at Self-Care in Patients With Multimorbidity: □ Cross-Sectional □ Usability Study. | 30 | 2024 | Cross-Sectional Usability Study. | '38 agreed to participate in the usability study. 30 patients completed the usability study.<br>The median age of the 38 participants was 75 (IQR 72.0-79.0) years. There was a slight majority of women (20/38, 52.6%) and the participants had a median of 8 (IQR 7.0-11.0) | Cognitive impairment, Parkinson disease, or cardiovascular disease. | 'To analyze the usability of a technological tool dedicated to health and self-care in patients with multimorbidity in primary care.' | Spain | 'Although usability effectiveness was high, the poor efficiency and usability satisfaction scores suggest that there are other factors that may interfere with the results.' |
|  |  |  |  | chronic diseases.’ |  |  |  |  |
| Monahan, et al.,<br>Development and Feasibility of SymTrak, a Multi-domain Tool for Monitoring Symptoms of Older Adults in Primary Care. | 31 | 2019 | Development and feasibility study. | ‘Phase 1: development of symtrack.<br>Four focus groups were held with 13 physicians (2 to 5 per group). Two focus groups were held with 13 advanced practice nurses (6 and 7 per group) Eight focus groups were held with 1 to 4 patients per group and eight separate focus groups were held with 1 to 4 caregivers per group, for a total of 16 dyads.<br>Phase 2: feasibility pilot with 81 participants (27 patients, their 27 caregivers, and 27 patients without a caregiver).’ | Does not state, but mental some mental health excluded. | ‘Develop and assess usability, administration time, and internal reliability of Symtrack – a digital tool for tracking symptoms in those with MLTC.’ | USA | ‘SymTrak demonstrates content validity, positive qualitative findings, high perceived usability, brief self-administered completion time, and good internal reliability.’ |
| Moran, et al.,<br>Investigating the needs and concerns of older adults with multimorbidity and their healthcare professionals for conceivable digital psychotherapeutic interventions. | 32 | 2022 | Qualitative study, one-to-one interviews. | ‘Participants were 11 older adults with multimorbidity (4 males; mean age: M =72.7 years) and 14 healthcare professionals including five clinical nurse specialists, four pharmacists, two general practitioners, one occupational therapist, one speech and language therapist and one dietician.' | Diabetes, COPD, chronic heart disease (CHD), (congestive heart failure) and others. | ‘To explore insights are into their specific concerns and issues around condition management and digital applications.’ | Republic of Ireland | ‘Key themes: patient feelings of anxiety or worry leading to an unwillingness to access essential information; the various mental health challenges faced by those with multimorbidity; the importance of personal values in providing motivation; and the importance of social support.' |
| Nambisan, et al.,<br>A Comprehensive Digital Self-care Support System for Older Adults With Multiple Chronic Conditions: Development, Feasibility, and Usability Testing of myHESTIA. | 33 | 2023 | Feasibility study. | 'Forty-five older patients, mixed ethnicities with MLTC + 15 geriatric clinicians and 10 carers across phase 1 and 2.' | Thirty-six different conditions. | 'To assess the efficacy and usability of myHESTIA.' | USA | 'Phase-1 show the need for a CDSSS. Phase-2 results demonstrate interest among older adults in using such a CDSSS and Phase-3 findings show that older adults found the tracking component of the system easy to use for capturing daily inputs.' |
| Northwood et al.,<br>Use of an electronic wellness instrument in the integrated health and social care of older adults: a group concept mapping study. | 34 | 2024 | 'Group concept mapping, a participatory mixed-methods approach.' | 'Persons were eligible to participate if they were an older adult (≥ 65 years), a care partner of an older adult, or staff or leader of an organization providing services to older adults. A total of 41 participants completed the idea generation step. For the sorting and rating session, 25 participants completed the sorting activity, and 19 participants completed the rating activity for importance and 15 for community capacity.' | No specific conditions – multiple chronic conditions. | 'The aim of this study was to develop, interRAI Check Up Self Report.' | Canada | 'Participants contributed to a cluster map of ten action areas - engagement of older adults and care partners, instrument's ease of use, accessibility of the assessment process, person-centred process, training and education for providers, provider coordination, health information integration (most important cluster), health system decision support (least important) and quality improvement, and privacy and confidentiality.' |
| Pettus et al.,<br>Internet-Based Resources for Disease Self-Care Among Middle-Aged and Older Women with Chronic Conditions. | 35 | 2017 | Qualitative research – telephone survey and interviews. | ‘427 females aged 44+ with one or more chronic condition.’ | Heart disease, cancer, stroke, diabetes, arthritis, asthma, hypertension or high blood pressure, emphysema, chronic bronchitis, depression, and anxiety. | ‘To describe and analyse disease types and healthcare utilization associated with Internet use among middle-aged and older women with 1 or more chronic diseases and identify which online resources they use.’ | USA | ‘65% reported using the Internet, a significantly fewer number of older women reported using the Internet, a significantly larger proportion of non-Internet users reported needing help learning what to do to manage their health conditions and needing help learning how to care for their health conditions.’ |
| Runz-jorgesson et al.,<br>Perceived value of eHealth among people living with multimorbidity: a qualitative study. | 36 | 2017 | Qualitative, interview study. | ‘Ten adults living with MLTC, 6 males, 4 females, mean age 68 years, 5.5 chronic conditions on average.’ | Chronic obstructive pulmonary disease (COPD), heart disease, diabetes, or depression. | ‘To explore challenges related to multimorbidity and patients’ perspectives on eHealth.’ | Denmark | ‘Patient-perceived value of eHealth varied, depending on their burden of illness and treatment: those with a greater burden had more positive perceptions of eHealth, and expressed more intention to use it. Lower disease burden links with less worth seen of eHealth. eHealth should only be optional.’ |
| Samal et al.,<br>Health information technology to improve care for people with multiple chronic conditions. | 37 | 2021 | Scoping review. | ‘MEDLINE, CINAHL, PsycINFO, EMBASE, Compendex, and IEEE Xplore databases for studies published in English between 2010 and 2020. 44 were included.’ | Not listed. | ‘To review evidence of digital applications and self-management for MLTC.’ | International | ‘Despite a growing body of literature on health IT interventions or multicomponent interventions including a health IT component for chronic disease management, current evidence for applying health IT solutions to improve care for PLWMCC is limited.’ |
| Shaw et al.,<br>Mobile health devices: will patients actually use them? | 38 | 2016 | Feasibility study. | ‘Six adult patients, healthy versus unhealthy.’ | Obesity, diabetes, hyperlipidemia, and/or hypertension. | ‘To develop a technological infrastructure to monitor biomarkers of those with MLTC.' | USA | ‘Device adherence declined across all participants during the study. Patients with chronic illnesses, with arguably the most to benefit from advanced (or increased) monitoring, may be less likely to adopt and use these devices compared to healthy individuals.' |
| Sheng, et al.,<br>Home-based digital health technologies for older adults to self-manage multiple chronic conditions: A data-informed analysis of user engagement from a longitudinal trial. | 39 | 2024 | A proof-of-concept trial with an action research design and mixed methods approach. | 'This paper reports on data from 60 Irish older adults with multiple chronic health conditions ($\geq 2$ of the following: COPD, HF, HD, and diabetes).' | Not listed. | 'The aim of this study is to assess the usability of ProACT.' | Republic of Ireland | '80% of participants using the technology devices for over 200 days. The submission frequency for symptom parameters (e.g. blood glucose (BG), blood pressure (BP), etc.) was highest. The majority of interactions happened in the morning time.' |
| Steele et al.,<br>Tying eHealth Applications to Patient Needs: Exploring the Use of eHealth for Community-Dwelling Patients With Complex Chronic Disease and Disability. | 40 | 2014 | Focus groups. | 'There were 14 total participants in the focus groups. The average age of participants was 64.4 years; NINE participants, all female.' | Not listed. | 'To determine the perceived needs of MLTC patients in relation to eHealth applications.' | Canada | Participants identified a need for open two-way communication and dialogue between themselves and their providers, Participants expressed common concerns around privacy and data security, accessibility, the loss of necessary visits, increased social isolation, provider burden, downloading responsibility onto patients for care management, entry errors, training requirements, and potentially confusing interfaces.' |
| Trinh, et al.,<br>Understanding older adults' motivations to use digital health portals. | 41 | 2024 | Qualitative study using interviews and questionnaires/surveys. | Twenty-six older adults (aged 60 to 78, M = 68.54) participated. | No specific conditions – 'chronic illnesses'. | 'To understand how older adults perceive digital health portals (DHPs), what motivates them to use them, and how their experience can be improved.' | USA | 'All the older adults shared that they were willing to communicate with their healthcare provider, and 25 of the 26 reported that their DHP supported their health needs. Friends and family supported general usage and provided additional guidance for DHP features.' |
| Wicks et al.,<br>Sharing health data for better outcomes on PatientsLikeMe. | 42 | 2010 | Cross-sectional survey. | 1,323 users from the Patients like me platform. | Amyotrophic lateral sclerosis [ALS], Multiple Sclerosis [MS], Parkinson's Disease, human immunodeficiency virus [HIV], fibromyalgia, and mood disorders). | 'Sought to describe the potential benefits of PatientsLikeMe.' | USA | 'Users perceived the greatest benefit in learning about a symptom they had experience. Other effects were better understanding of the side effects of their treatments, finding another patient who had helped, help with decisions to around medication and increasing levels of comfort in sharing personal health information among those who had initially been uncomfortable. Analysis of the Web access logs showed that participants who used more features of the site (e.g., posted in the online forum) perceived greater benefit.' |
| Wu, et al.<br>Associations between e-health literacy and chronic disease self-management in older Chinese patients with chronic non-communicable diseases: a mediation analysis. | 43 | 2022 | Cross-sectional study of randomly selected patients. | '300 older patients >60 with Chronic non-communicable diseases (CNCDs) from four nursing homes in Hunan province, China, from July to November 2021. Patients with mental disorders, communication impairment, or serious illnesses and complications were excluded.' | No specific conditions - Chronic non-communicable diseases. | 'To explore the factors that influence chronic disease self-management (CDSM) and verify self-efficacy as the mediator between e-health literacy and self-management behavior in older patients with CNCDs.' | China | 'Smart medical devices were positively correlated with CDSM in older adults, which agrees with previous findings. Medical professionals can give personalized advice and specialized guidance to increase product use and benefit rate. Devices should focus on simplicity of operation, functional variety, efficiency, and practicality. Self-efficacy plays a partial intermediary role between e-health literacy and self-management in older patients with CNCDS.’ |
| Xu, et al.,<br>Use, satisfaction, and preference of online health services among older adults with multimorbidity in Hong Kong primary care during COVID-19. | 44 |  | Cross-sectional survey. | 752 older people aged 55 or over with multimorbidity. | Not listed. | ‘To explore the use of social media in older adults with multimorbidity in Hong Kong.’ | Hong Kong | ‘Lower education levels, fewer internet connection issues, and more self-efficacy on mobile applications were associated with a higher level of online satisfaction after adjustment. Fewer internet connection issues and more self-efficacy on mobile applications were associated with participants’ preference for online services.’ |
| Yoo et al.,<br>A Ubiquitous Chronic Disease Care system using cellular phones and the internet. | 45 | 2009 | RCT. | 111 participants:<br>- Control group ($n = 54$).<br>- Intervention group ($n = 57$).<br>Aged between 30 and 70 years of age. | Type 2 diabetes and hypertension, particularly in overweight people. | ‘To assess the effectiveness and applicability of the Ubiquitous Chronic Disease Care (UCDC).’ | South Korea | ‘After 12 weeks, there were significant improvements in HbA(1c) in the intervention group, a significant reduction in systolic and diastolic blood pressure, as well as improvements in total cholesterol, low-density lipoprotein-cholesterol and triglyceride levels in the intervention group. There was a significant increase in adiponectin levels in the intervention group.’ |
| Zulman et al.,<br>How Can eHealth Technology Address Challenges Related to Multimorbidity? Perspectives from Patients with Multiple Chronic Conditions. | 46 | 2015 | Qualitative research including:10 focus groups, three to eight participants in each. | ‘Fifty-three individuals with $\geq 3$ chronic conditions and experience using technology for health- related purposes.’ | High blood pressure, chronic pain, arthritis, rheumatism, depression, diabetes, migraines, PTSD, lung issues, cancer, prostate problems. | ‘To understand self-management and healthcare opportunities to support patients through new and enhanced eHealth technology.’ | USA | ‘Participants most frequently use digital applications to search for health information (96 %), communicate with health care providers (92 %), track medical information (83 %), track medications (77 %), and support decision-making about treatment (55 %).<br>Patients manage a high volume of information, visits, and self-care tasks; they need to coordinate, synthesize, and reconcile health information from multiple providers and about different conditions; their unique position at the hub of multiple health issues requires self-advocacy and expertise.’ |

\*The text included in this table has largely been extracted directly from the relevant papers.

**Supplementary Table 4:**
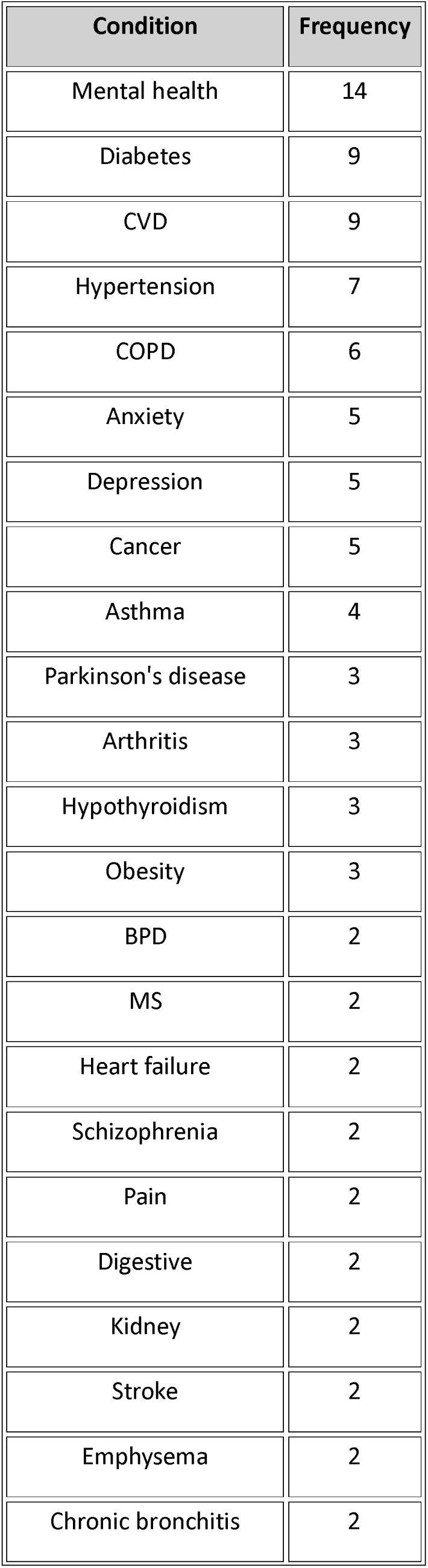
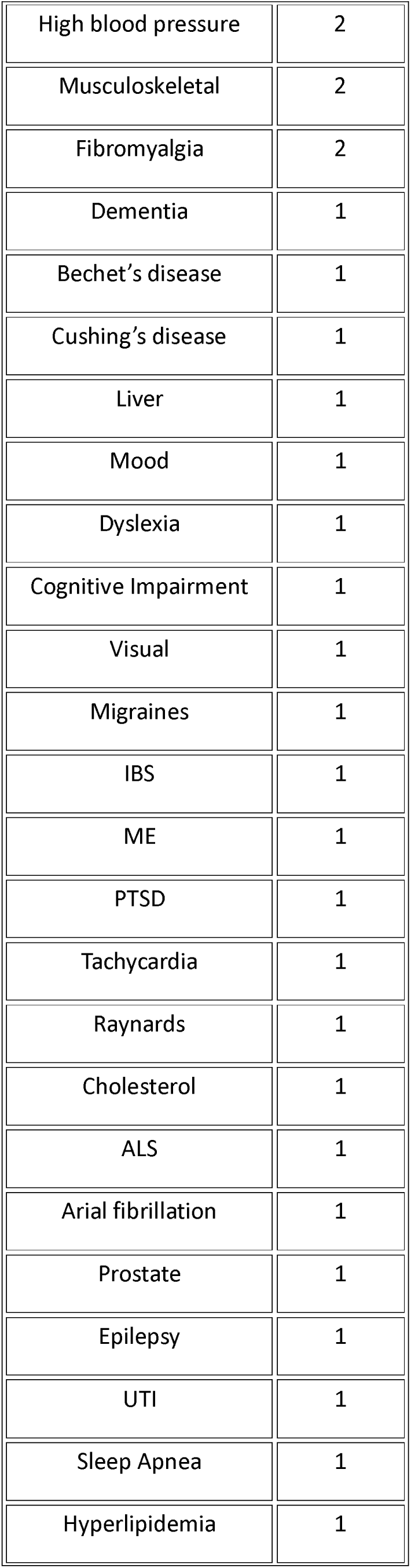
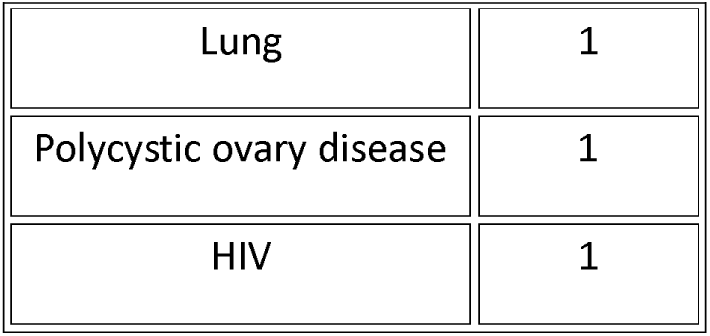
51 chronic conditions referred to in the studies and frequency.

| Condition | Frequency |
| --- | --- |
| Mental health | 14 |
| Diabetes | 9 |
| CVD | 9 |
| Hypertension | 7 |
| COPD | 6 |
| Anxiety | 5 |
| Depression | 5 |
| Cancer | 5 |
| Asthma | 4 |
| Parkinson's disease | 3 |
| Arthritis | 3 |
| Hypothyroidism | 3 |
| Obesity | 3 |
| BPD | 2 |
| MS | 2 |
| Heart failure | 2 |
| Schizophrenia | 2 |
| Pain | 2 |
| Digestive | 2 |
| Kidney | 2 |
| Stroke | 2 |
| Emphysema | 2 |
| Chronic bronchitis | 2 |
| High blood pressure | 2 |
| Musculoskeletal | 2 |
| Fibromyalgia | 2 |
| Dementia | 1 |
| Bechet's disease | 1 |
| Cushing's disease | 1 |
| Liver | 1 |
| Mood | 1 |
| Dyslexia | 1 |
| Cognitive Impairment | 1 |
| Visual | 1 |
| Migraines | 1 |
| IBS | 1 |
| ME | 1 |
| PTSD | 1 |
| Tachycardia | 1 |
| Raynards | 1 |
| Cholesterol | 1 |
| ALS | 1 |
| Arial fibrillation | 1 |
| Prostate | 1 |
| Epilepsy | 1 |
| UTI | 1 |
| Sleep Apnea | 1 |
| Hyperlipidemia | 1 |
| Lung | 1 |
| Polycystic ovary disease | 1 |
| HIV | 1 |

